# 222 nm Far-UVC from filtered Krypton-Chloride excimer lamps does not cause eye irritation when deployed in a simulated office environment

**DOI:** 10.1101/2022.11.26.22282779

**Authors:** Obaid Kousha, Paul O’Mahoney, Robert Hammond, Kenneth Wood, Ewan Eadie

## Abstract

Far-UVC, in the form of filtered Krypton-Chloride lamps, is a promising technology for reducing airborne transmission of disease. Whilst significant research has been undertaken to investigate skin safety of these lamps, less work has been undertaken on eye safety. In particular, there is very limited data on human eye safety or discomfort from the deployment of this germicidal technology. In this pilot study, immediate and delayed eye discomfort were assessed in a simulated office environment with deployment of Krypton-Chloride lamps. The discomfort was assessed immediately post-exposure to the Far-UVC and several days after exposure using the validated, standardized Standard Patient Evaluation Eye Dryness (SPEED) and Ocular Surface Disease Index (OSDI) questionnaires. Our results show that there was no significant eye discomfort or adverse effects from the deployment of Far-UVC in this simulated office environment, even when the lamps were operated continuously. In addition, through collection of bacteria and fungi on agar plates, with this non-optimised lamp arrangement a statistically significant reduction in pathogens of 52% was observed. Far-UVC in this simulated office environment did not cause any clinically significant eye discomfort and was effective at reducing pathogens in the room. These results contribute an important step to further investigation of the interaction of Far-UVC with the human eye.

## Introduction

Transmission of airborne pathogens, such as severe acute respiratory syndrome coronavirus 2 (SARS-CoV-2), influenza, measles and tuberculosis, have crucial global implication. The coronavirus disease 2019 (COVID-19) pandemic, which is caused by SARS-CoV-2, highlighted the short-term and long-term devastating effects that these pathogens can have on all facets of human life including health, education, and economy.

The risk of transmission of airborne pathogens increases in poorly ventilated indoor spaces where groups of people gather (1). Examples of these environments include educational institutions such as schools and universities, healthcare facilities, offices, public transport hubs, retail and commercial areas. To reduce transmission, 254 nm germicidal ultraviolet (GUV) has been utilized in the past to good effect(2–4). However, the major challenge of using the conventional 254nm GUV is that accidental over-exposure of human skin or eye causes potentially painful sunburn-type reaction(5).

Far-UVC, a germicidal ultraviolet-C radiation with typical wavelength ranging between 200 nm and 230 nm, can potentially be used to meet this challenge. Filtered Krypton Chloride (KrCl) excimer lamps with a primary emission wavelength of 222nm and low residual emission of other ultraviolet wavelengths is a common source of Far-UVC(6). Far-UVC has been shown to inactivate a range of pathogens, including bacteria and viruses, in laboratory settings(7–12). It has also been shown to effectively inactivate aerosolized bacteria in a room-sized chamber(13).

Due to a limited penetration depth of 222 nm in tissue, there are no acute effects observed in skin exposed to a KrCl lamp, when it is filtered to minimise longer wavelength emissions(14–16). There is also evidence suggesting that the induction of non-melanoma skin cancer is unlikely at current exposure limits(17, 18), although other potential long term effects need to be explored.

Eye safety of Far-UVC has mostly been studied in animal models thus far. Kaidzu et al. demonstrated in rats that even at exposure doses of 600 mJcm^-2^, the corneal surface integrity is maintained, which is a surrogate marker of corneal irritation(19). Furthermore, in mice, rat, rabbit and porcine eyes, the 222 nm appears to only significantly penetrate the corneal epithelium at exposures above 1500 mJcm^-2^ (20). The latest work from the same group showed that corneal limbal stem cells are also safe from damage at 600 mJcm^-2^ exposure in rat and porcine models, as shown by the absence of cyclobutane pyrimidine dimer formation in the stem cells and the subsequent normal function of the stem cells post irritation demonstrated by normal turnover of the corneal epithelium(21).

Data on the effect of Far-UVC on human eyes is limited. In one study, where three individuals were exposed to 220 nm produced by an irradiation monochromator with Xenon-Mercury high pressure lamp, the threshold before photokeratitis developed was determined to be 10 mJcm^-2^, delivered in 256 seconds with participants staring directly at the source(22). However in a typical deployment of Far-UVC, room occupants are unlikely to be subject to direct irradiation of the eye from down-welling Far-UVC lamp installations. It has also been argued that the 10 mJcm^-2^ is incorrect due to the bandwidth of the monochromator used by Pitts in 1973 and the true threshold may be much higher than noted(20, 23). In more recent work on Far-UVC, six ophthalmologists exposed to filtered 222 nm Far-UVC in their work environment for more than one year, demonstrated no ill eye effects(24).

The aim of this pilot study was to investigate in a systematic manner whether filtered Krypton Chloride lamps at two different exposure levels, when deployed in an office type environment, induce any eye irritation.

## Materials and Methods

### Research Ethics

This work was approved by the University of St Andrews Teaching and Ethics Committee (Approval code: MD15737) and adheres to the tenets of the declaration of Helsinki.

### Study Design

Participants were recruited from the University of St Andrews and surrounding areas through email advertisement and word of mouth. Potential participants who responded to the recruitment were screened by one of the investigators (OK). The exclusion criteria included anyone with a pre-existing diagnosed eye condition, anyone who may be photosensitive, anyone taking medication or herbal supplements that could induce photosensitivity, anyone who was immunosuppressed or had a history of skin cancer and anyone who could not understand or comply with the protocol requirements, timetables, instructions and protocol-stated restrictions. A participant information sheet was provided to the potential participants and written informed consent was taken more than 24 hours later. Participants were reminded that they could withdraw from the study at any time and without any penalty.

The participants were allocated into one of five groups (A - E) depending on the date they responded to the advert and the dates they were available to participate. Participants were not randomised to the groups by any criteria but self-selected their group based on their availability and convenience. Each group participated for a total of 3 days in the study and each study day was separated by a minimum of 3 days. During a study day, the participants were instructed to remain in an adapted classroom at the University of St Andrews from ten o’clock in the morning to four o’clock in the afternoon. If they wore contact lenses or glasses on the first day of the study, they had to do the same for all the remaining days of the study. At least one of the investigators were present during this time to supervise the study day. Whilst in the classroom the participants were free to undertake tasks as they wished, for example, work on a laptop, read a book, etc. A 15-minute break in the morning, 30-minute lunch break and 15-minute break in the afternoon were provided. Leaving the room at other times was prohibited, except to use the toilet. This design meant the participants spent a minimum of 5 hours in the classroom on each study day.

### Far-UVC Lamps

Eight filtered krypton-chloride excimer lamps (Biotile, Biocare UV, Warrington, United Kingdom) were installed in the ceiling of the classroom prior to the recruitment process. The location of the lamps within the room is shown in Figure 1 and the spectral emission of the lamps is given in Supplementary Figure 1. The room has dimensions 12 m x 5.9 m x 3 m and was arranged as shown in Figure 1. As previously described, the room has mechanical ventilation with four air inlets and three open windows for outlets providing a ventilation rate of 6.8 air-changes-per-hour (ACH)(25).

**Figure 1.**
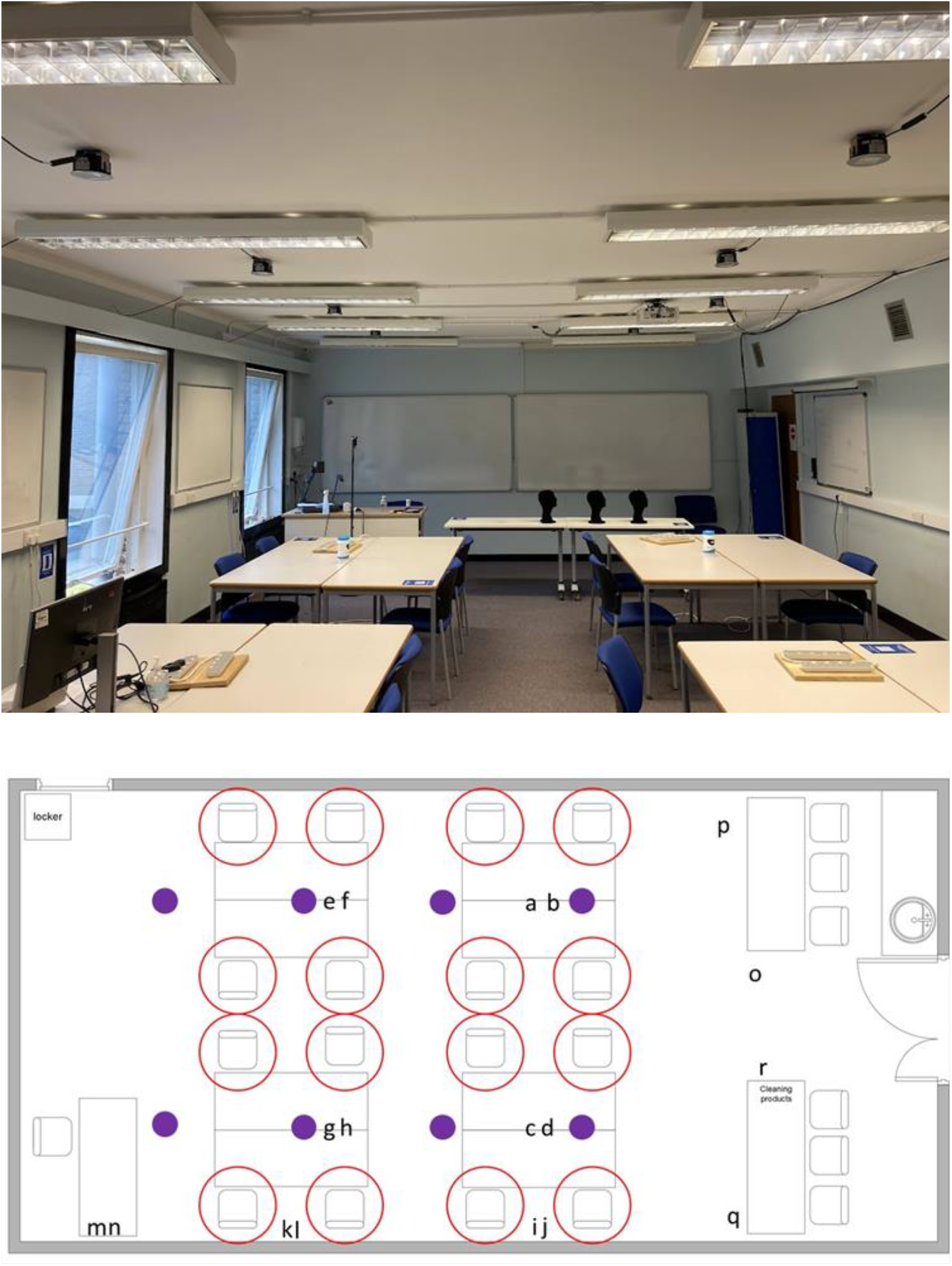
(Top) A photograph of the classroom used during the study. The KrCl excimer lamps can be seen secured to the ceiling between the normal room lighting. (Bottom) A schematic of the classroom with the location of the KrCl excimer lamps indicated by purple dots. The available seating positions for the participants are indicated with red circles and the locations of the agar plates for microbiological sampling are indicated by the letters a – r.

Each study group experienced three different Far-UVC exposures during the study. The exposures were

- <u>NO</u> Far-UVC Lamps switched on
- Far-UVC Lamps on <u>ALL</u> the time
- Far-UVC Lamps with a <u>DUTY</u> cycle of 30 seconds on, 270 seconds off (1:9).

The three scenarios were randomly assigned to study day 1, 2 or 3. Participants were not informed of the Far-UVC exposure scenario on each study day. However, there was also no attempt to disguise the Far-UVC exposure scenario and therefore, for the purposes of study design, the participants were not regarded as blinded. To provide an indication of the Far-UVC dose received by participants each wore a brimless cloth cap with UVC Dosimeters (UVC 222 Dots, Intellego Technologies, Solna, Sweden) fixed to the top. At the end of each study day the dosimeters were visually assessed and compared to the included reference chart which provided a dose range, e.g., 0, 0-20, 20-50 mJ cm^-2^, dependent on the resultant colour of the dosimeter.

### Eye Discomfort

Eye and visual discomfort were ascertained by asking the participants to complete validated questionnaires. The Standard Patient Evaluation Eye Dryness (SPEED) questionnaire was completed by participants at the start of each study day, at the end of each study day and the day after each study day(26). The SPEED questionnaire relates to symptoms which are currently being experienced and provided an indication of any immediate discomfort as a result of Far-UVC exposure. Questionnaire scores were calculated and changes from baseline (start of each study day) were compared for the three different Far-UVC exposures.

Another questionnaire, the Ocular Surface Disease Index (OSDI) questionnaire, was completed by participants at the start of each study day and one week after the last study day(27). The OSDI questionnaire provides an indication of discomfort felt over the previous week and therefore provided an indication of any delayed effects from the Far-UVC exposure. A similar analysis to the SPEED questionnaire scoring was performed comparing each Far-UVC exposure to their OSDI score at the start of the study.

### Microbiology

Whilst the purpose of this study was to investigate the potential for eye discomfort during typical office-based Far-UVC exposure, the opportunity was used to also acquire microbiological data. 18 agar plates were placed at various locations throughout the room between 2 pm and 4 pm on some of the study days (Figure 1). The locations were broadly grouped into table top (a – h), window ledge (i – n) and floor (o – r).

After 2 hours exposure the permissive agar plates (BHI agar, Sigma, UK) were collected and incubated at 30°C for 48 hours and then 37°C for 24 hours. After the 30°C incubation colonies were identified & counted and the plates photographed. After the 37°C incubation any further colonies were identified & counted. Data was recorded by blinded operators.

### Statistical Analysis

Statistical analysis of the SPEED, OSDI and Microbiology data was performed using GraphPad Prism 9.4.1 (GraphPad Software LLC, San Diego, USA).

## Results

### Demographics of participants

38 participants were recruited for the study with one failing to attend the first study day. Of the remaining 37 participants, 46% (n=17) were female. The mean age was 34 years with standard deviation of 14.5 years and a range of 18 years to 68 years. One participant wore contact lenses during the study (3%) and 20 wore glasses (55%) - 10 glasses for distance (27%), eight reading glasses (22%) and two wore varifocals (6%). During the study days, the majority of participants sat and undertook tasks on laptops, with a small number reading a book or watching a tablet. Although it was not a requirement to sit in the same location on each study day, most of the participants did. The study scenarios for each group are described in table 1.

**Table 1.** Far-UVC exposure scenarios for each of the study groups A – E. NO indicates a day when there was no Far-UVC exposure, DUTY a day when a duty cycle of 30 seconds on and 270 seconds off for the KrCl excimer lamps was deployed and ALL when the KrCl excimer lamps were continuously emitting for the full day.

| Group (No. of participants) | Study Day 1 | Study Day 2 | Study Day 3 |
| --- | --- | --- | --- |
| **A (8)** | NO | DUTY | ALL |
| **B (4)** | DUTY | ALL | NO |
| **C (7)** | ALL | NO | DUTY |
| **D (10)** | DUTY | ALL | NO |
| **E (8)** | NO | ALL | DUTY |

### Far-UVC Exposure

Directly under the lamp, at a height of two meters from the ground, the measured irradiance was 11.5 μWcm^-2^. At this point in space, a 6-hour exposure would result in a UV dose of 248 mJcm^-2^ with the lamps on continuously. With the lamps on a duty cycle as indicated previously, there would be 72 cycles within 6 hours which would result in a UV dose of 24.8 mJcm^-2^.

None of the participants stood directly under a lamp for a 6-hour period. The majority of participants sat down during the study. On study days where there were no Far-UVC lamps switched on, no UV dose was recorded on the UVC dosimeters. On DUTY days the majority of participants received no UV dose (66%) with 34% receiving a dose between 0 and 20 mJcm^-2^. When the lamps were on continuously (ALL), 62% of participants received a dose between 0 and 20 mJcm^-2^, 25% between 20 and 50 mJcm^-2^ and 13% received no UV dose (Figure 2). Due to the nature of the UVC dosimeters it was not possible to more accurately define the UV exposure.

**Figure 2.**
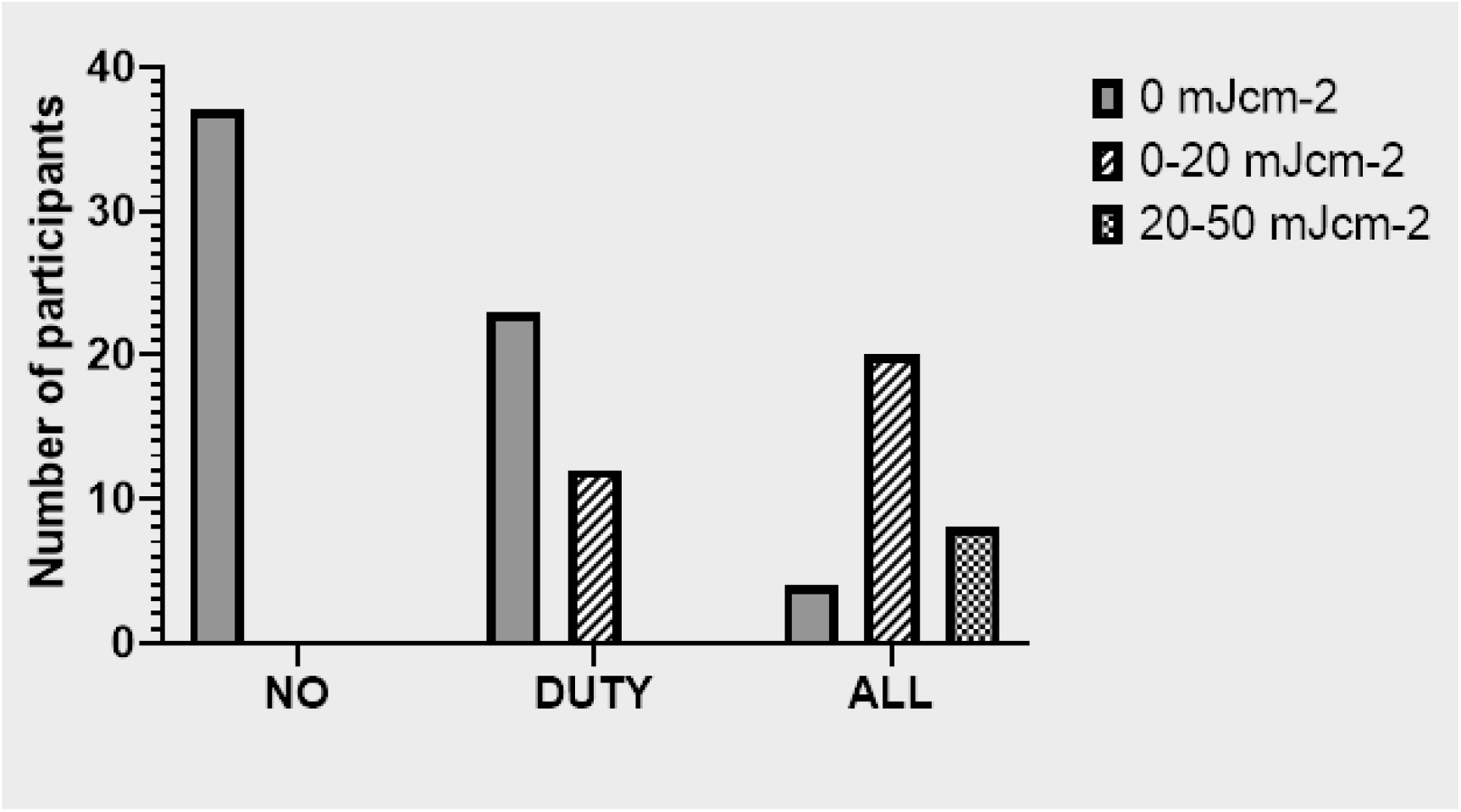
Number of participants and associated Far-UVC dose for each of the three exposure scenarios (NO, DUTY, ALL) as recorded by UVC Dosimeters (UVC 222 Dots, Intellego Technologies, Solna, Sweden) affixed to the top of brimless cloth caps worn by participants.

### Eye Discomfort

The mean SPEED score of the participants is given in Table 2. There was no evidence that the mean SPEED score was different at the start of the day, end of the day or the following day for either the NO, DUTY or ALL exposure. There was also no statistically significant difference in the SPEED score when compared to the baseline questionnaire score at the start of the day (Friedman Test). There was no evidence in this study of immediate eye discomfort as a result of the Far-UVC being deployed. Further data exploration indicated that there was also no association between those who received the highest UV dose or those who wore glasses and the SPEED score. However, the participant numbers were too small in this sub-analysis to apply any statistical tests.

**Table 2.**
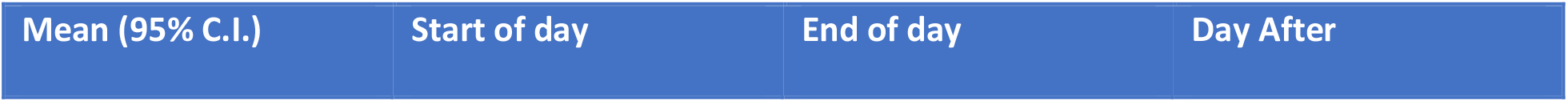
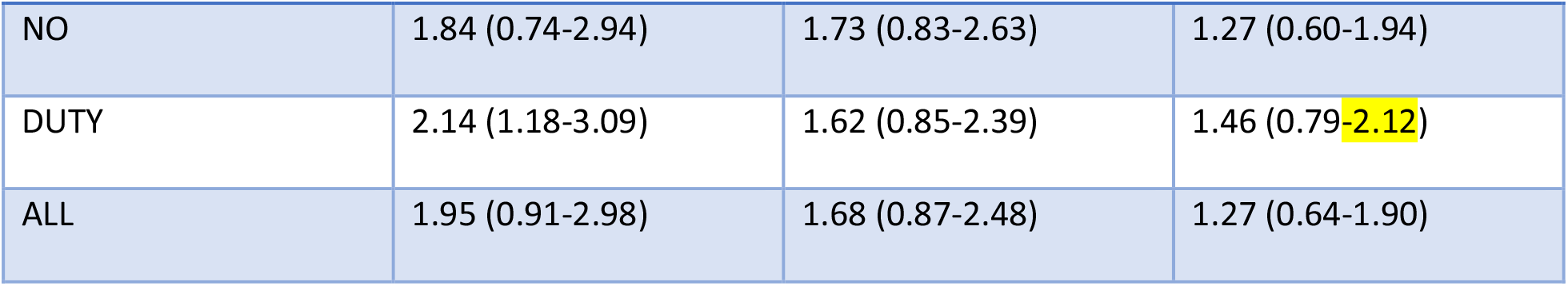
Mean SPEED questionnaire score (95% confidence interval) at the start of the study day, end of study day and the day after the study day for each of the three exposure scenarios (NO/DUTY/ALL).

| Mean (95% C.I.) | Start of day | End of day | Day After |
| --- | --- | --- | --- |
| NO | 1.84 (0.74-2.94) | 1.73 (0.83-2.63) | 1.27 (0.60-1.94) |
| DUTY | 2.14 (1.18-3.09) | 1.62 (0.85-2.39) | 1.46 (0.79-2.12) |
| ALL | 1.95 (0.91-2.98) | 1.68 (0.87-2.48) | 1.27 (0.64-1.90) |

Similar to the SPEED score, there was no evidence that the OSDI scores after each exposure day were different from the baseline, pre-study OSDI scores or different from each other (Figure 3). Only one of the OSDI scores was outside the normal range of 0-12 and this result occurred pre-study. The mean OSDI scores were 2.7 at the start of the study (PRE), 1.4 following no exposure (NO), 1.7 following the study day with duty cycle exposure (DUTY) and 1.4 following the study day with continuous lamp exposure (ALL). As with the SPEED score, there was no association between those who received the highest UV dose or those who wore glasses and the OSDI score.

**Figure 3.**
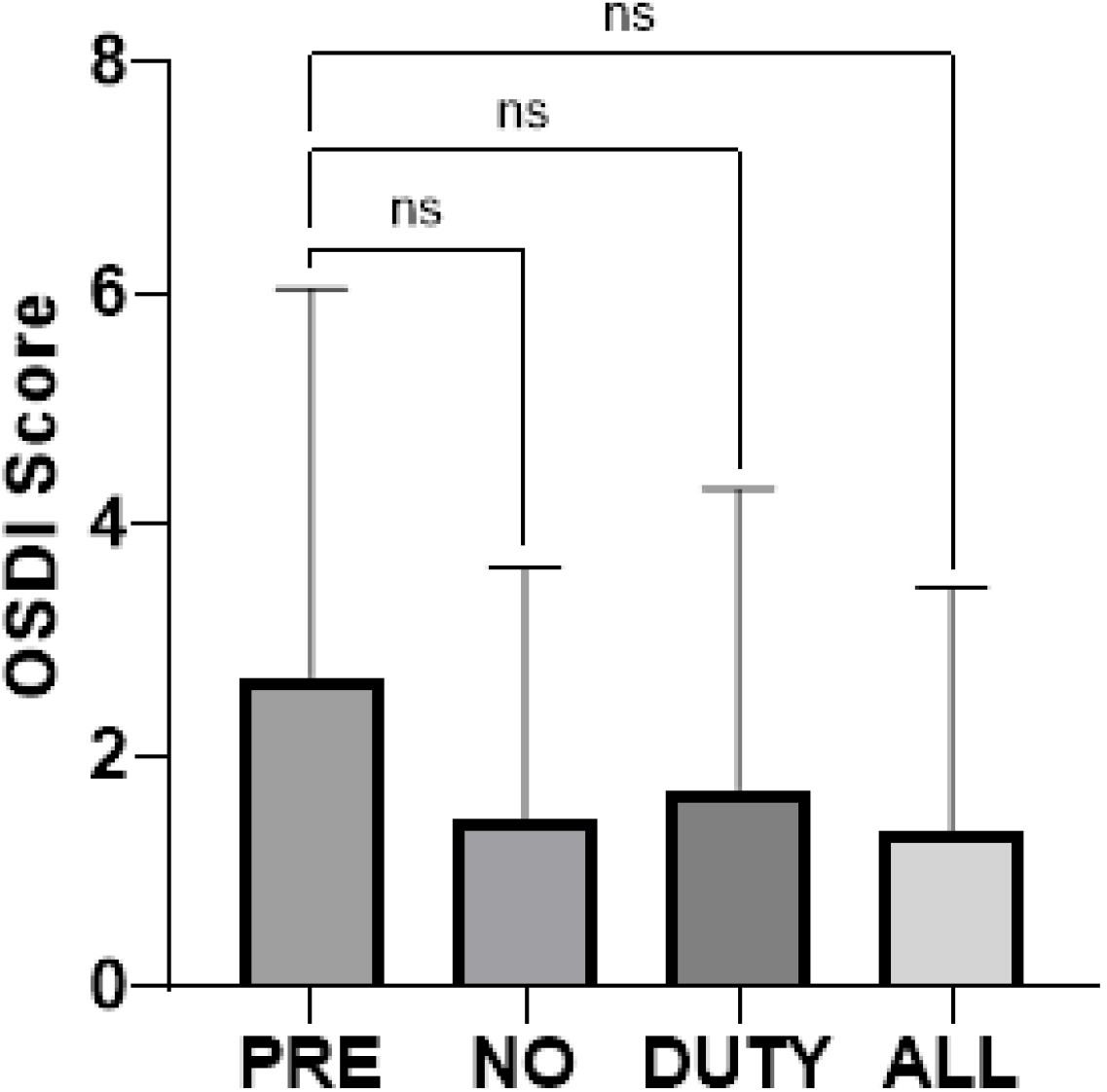
OSDI scores for pre-study, following NO exposure, DUTY cycle exposure and ALL exposure. Columns represent the mean OSDI score with lines representing the standard deviation. There is no statistically significant difference in the OSDI scores for any exposure scenario (Friedman test).

**Figure 5.**
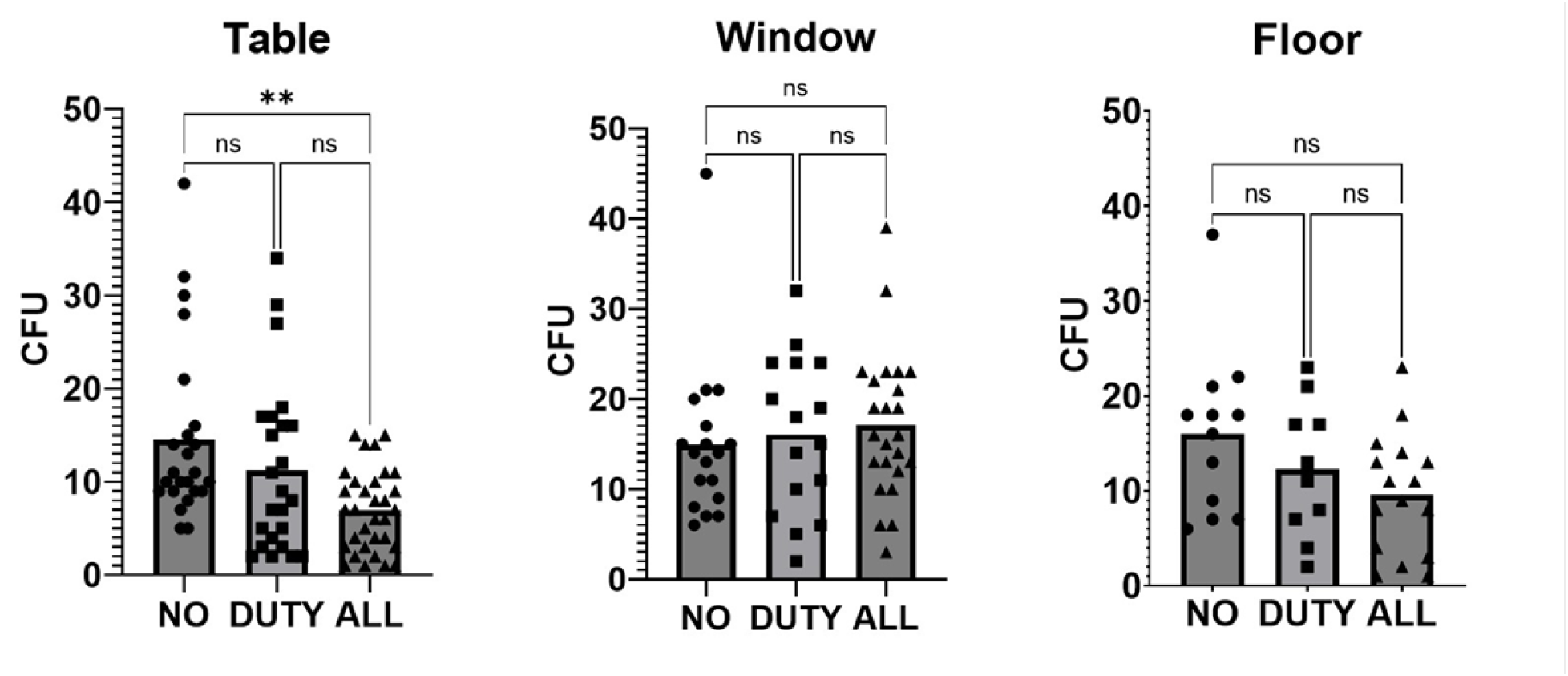
Number of colony forming units (CFU) counted on agar plates for each of the exposure scenarios (NO/DUTY/ALL). Individual data points are plotted on the graphs along with the mean number of CFU (column). Data are grouped for the agar plates located on the tables (left), window ledges (middle) and floor (right). Data from the tables and the floor agar plates show a general trend for lower colony forming units during Far-UVC exposure however only the table data shows a statistically significant difference between the no exposure scenario and the lamps on continuously. A lack of data points may be the reason for not reaching statistical significance or the differences seen could be due to chance.

### Adverse Events

During the study, one participant reported to develop a non-infected chalazion on the lower eyelid of the left eye a week after their first study day. Since their first day was NO exposure, it was not believed that this adverse event was as a result of the study.

### Microbiology

The only statistically significant difference in CFU was from the agar plates on the tables, between the NO exposure day and the ALL exposure day, with mean CFU of 14.5 and 6.9 respectively providing an average reduction in CFU of 52% (Ordinary One-Way Anova). A mean reduction in CFU of 22% (mean CFU 11.3) was observed in the data from the tables between the NO exposure and DUTY cycle exposure days but this was not statistically significant.

The data from the sample plates on the floor showed a mean reduction from the NO exposure day (16.0 CFU) of 23% (12.3 CFU) and 40% (9.6 CFU) for the DUTY cycle and ALL exposure scenarios respectively. The data from the agar plates on the window ledges showed a 7.5% and 14.3% increase when comparing DUTY cycle and ALL exposure to the NO exposure day (no exposure = 14.9 CFU, duty cycle exposure = 16.1 CFU and continuous exposure = 17.1 CFU). None of these reductions or increases was statistically significant

## Discussion

This study has assessed, in a controlled and standardized manner, self-reported eye discomfort when filtered Krypton-Chloride lamps are deployed in a typical office/classroom environment using a validated standardised questionnaire. The results indicate that there is no clinically significant immediate or delayed discomfort experienced when Far-UVC is deployed as described.

Moreover, eye symptoms were monitored with repeated administration of OSDI questionnaire for the duration of the study in each participant without any worsening symptoms. By administering the SPEED questionnaire before, immediately after, and 24 hours after exposure it was ensured to detect any delayed discomfort. Importantly, one of the major potential confounders, i.e. wearing glasses or contact lenses did not have any effect on the development of eye symptoms since individuals who didn’t wear refractive correction aids remained symptom free. In addition, the self-reported eye scores of individuals with higher maximum exposure were no different from individuals with zero exposure as per dosimeter indicator. In all groups the self-report eye scores didn’t change in a statistically significant way from the baseline.

One historical study has reported eye irritation after 10mJcm^-2^ of 220nm FAR-UVC exposure but was potentially flawed in its design due to bandwidth of the irradiation monochromator used in the study(23).

Although we measured the Far-UVC exposure to the top of the head, we cannot know the actual eye exposure of our participants. The relationship between Far-UVC exposure to the top of the head and the eye is highly variable and will depend upon multiple factors, including distance from and angle to the lamps throughout the exposure duration. Duncan *et al*. demonstrated in a mannequin study that the eye received, on average, 5.8% of the dose measured from the top of the head. However the variability in recorded measurements was very large, including several measurements where the eye received no UV dose despite significant exposure on the top of the head(28). Similar variability was observed in a study by First *et al*., which found variation in participant eye dose of between 3% and 37% compared to a calculated dose(29). We can therefore conclude that whilst we do not know the exact eye exposures of our participants, it is unlikely that eye exposure will have been higher than the recorded top of head exposure, and could have been just a few percent or less of this measurement. As such our study should not be regarded as a defining exposure level study but more a representation of what may be experienced in typical deployment of Far-UVC.

Our study is the largest human study to date that evaluates eye discomfort effects of Far-UVC when deployed in a simulated real-world environment. Although formal sample size was not carried out in this study, this seminal work can pave the way for an appropriately powered study designed to ascertain more definitively whether eye irritation occurs due to Far-UVC exposure. Given the avant-garde nature of this work, we restricted participants to individuals with healthy eyes. This limits the applicability of this work to individuals with already compromised ocular surface. Indeed, exposing individuals with ocular surface disease to Far-UVC will require careful consideration of interaction of the Far-UVC with diseased eye. This work does not evaluate the cumulative effects of repeated Far-UVC exposure without a washout period on eye irritation over a longer time, which could potentially be the case if this technology is deployed in public spaces.

Thus far, most animal studies have investigated the effect of Far-UVC on the different layers of the cornea and its penetration potential into the deeper ocular tissues. There is one study investigating the health of corneal limbal stem cells in animal models. Investigating the structure and function of the limbal stem cells will be crucial in determining if Far-UVC will have ill effect on the cornea in the long run. There are no reports of the effect of Far-UVC on conjunctival stem cells in literature. Studying these two populations of stem cells will ascertain carcinogenic potential of Far-UVC in ocular surface.

Additionally, potentially non-carcinogenic effects of the Far-UVC on the ocular surface including its interaction with tear film layer, effects on the conjunctival cells such as goblet cells, and long-term fibrogenic properties of Far-UVC in causing conditions such as pterygium need to be elucidated.

Whilst this study was not designed to investigate the inactivation of pathogens, it was encouraging to observe a reduction in sampled bacteria and fungi as the quantity of Far-UVC increased, even if some results were not statistically significant. Of particular interest were results from the agar plates placed on the floor of the room, which showed a reduction in colonies with Far-UVC. These plates were distant from direct Far-UVC irradiation, as shown in Figure 1, and therefore were representative of a reduction in circulating pathogen within the room when Far-UVC was deployed.

Therefore, whilst not definitive, this study does provide good preliminary evidence that acute eye discomfort is not experienced by individuals when filtered KrCl lamps are deployed in a real-world environment at intensity levels sufficient to reduce circulating pathogen load. Further study is warranted to expand the inclusion criteria and number of participants, and to determine the received ocular dose of Far-UVC.

## Data Availability

All data produced in the present study are available upon reasonable request to the authors

## Acknowledgements

We would like to thank Mike Humphreys, Adrian Leatherland and BiocareUV for the loan and installation of the filtered Krypton-Chloride lamps. We would also like to thank Intellego Technologies for the UVC dosimeters. The preceding individuals had no contribution to the conception, design, execution or analysis of this study.

## References

1. Miller, S. L., Nazaroff, W. W., Jimenez, J. L., Boerstra, A., Buonanno, G., Dancer, S. J., Kurnitski, J., Marr, L. C., Morawska, L. and Noakes, C. (2021) Transmission of SARS-CoV-2 by inhalation of respiratory aerosol in the Skagit Valley Chorale superspreading event. Indoor Air 31, 314–323. https://doi.org/10.1111/INA.12751.

2. Wells, W. F., Wells, M. W. and Wilder, T. S. (1942) The Environmental Control of Epidemic Contagion: I. An Epidemiologic Study of Radiant Disinfection of Air in Day Schools. Am. J. Epidemiol. 35, 97–121. https://doi.org/10.1093/oxfordjournals.aje.a118789.

3. Mphaphlele, M., Dharmadhikari, A. S., Jensen, P. A., Rudnick, S. N., Van Reenen, T. H., Pagano, M. A., Leuschner, W., Sears, T. A., Milonova, S. P., Van Der Walt, M., Stoltz, A. C., Weyer, K. and Nardell, E. A. (2015) Institutional tuberculosis transmission: Controlled trial of upper room ultraviolet air disinfection: A basis for new dosing guidelines. Am. J. Respir. Crit. Care Med. 192, 477–484. https://doi.org/10.1164/rccm.201501-0060OC.

4. Escombe, A. R., Moore, D. A. J., Gilman, R. H., Navincopa, M., Ticona, E., Mitchell, B., Noakes, C., Martínez, C., Sheen, P., Ramirez, R., Quino, W., Gonzalez, A., Friedland, J. S. and Evans, C. A. (2009) Upper-Room Ultraviolet Light and Negative Air Ionization to Prevent Tuberculosis Transmission. PLOS Med. 6, e1000043. https://doi.org/10.1371/journal.pmed.1000043.g001.

5. Oliver, H., Moseley, H., Ferguson, J. and Forsyth, A. (2005) Clustered outbreak of skin and eye complaints among catering staff. Occup. Med. (Chic. Ill). 55, 149–153. https://doi.org/10.1093/occmed/kqi021.

6. Barnard, I. R. M., Eadie, E. and Wood, K. (2020) Further evidence that far-UVC for disinfection is unlikely to cause erythema or pre-mutagenic DNA lesions in skin. Photodermatol. Photoimmunol. Photomed. 36, 476–477. https://doi.org/10.1111/phpp.12580.

7. Buonanno, M., Ponnaiya, B., Welch, D., Stanislauskas, M., Randers-Pehrson, G., Smilenov, L., Lowy, F. D., Owens, D. M. and Brenner, D. J. (2017) Germicidal Efficacy and Mammalian Skin Safety of 222-nm UV Light. Radiat. Res. 187, 493–501. https://doi.org/10.1667/rr0010cc.1.

8. Buonanno, M., Welch, D., Shuryak, I. and Brenner, D. J. (2020) Far-UVC light (222 nm) efficiently and safely inactivates airborne human coronaviruses. Sci. Rep. 10, 1–8. https://doi.org/10.1038/s41598-020-67211-2.

9. Welch, D., Buonanno, M., Shuryak, I., Randers-Pehrson, G., Spotnitz, H. M. and Brenner, D. J. (2018) Effect of far ultraviolet light emitted from ann optical diffuser on methicillin-resistant Staphylococcus aureus in vitro. PLoS One 13. https://doi.org/10.1371/journal.pone.0202275.

10. Narita, K., Asano, K., Naito, K., Ohashi, H., Sasaki, M., Morimoto, Y., Igarashi, T. and Nakane, A. (2020) Ultraviolet C light with wavelength of 222 nm inactivates a wide spectrum of microbial pathogens. J. Hosp. Infect. 105, 459–467. https://doi.org/10.1016/j.jhin.2020.03.030.

11. Ma, B., Gundy, P. M., Gerba, C. P., Sobsey, M. D. and Linden, K. G. (2021) UV Inactivation of SARS-CoV-2 across the UVC Spectrum: KrCl* Excimer, Mercury-Vapor, and Light-Emitting-Diode (LED) Sources. Appl. Environ. Microbiol. 87. https://doi.org/10.1128/AEM.01532-21.

12. Ma, B., Linden, Y. S., Gundy, P. M., Gerba, C. P., Sobsey, M. D. and Linden, K. G. (2021) Inactivation of coronaviruses and phage Phi6 from irradiation across UVC wavelengths. Environ. Sci. Technol. Lett. 8, 425–430. https://doi.org/10.1021/acs.estlett.1c00178.

13. Eadie, E., Hiwar, W., Fletcher, L., Tidswell, E., O’Mahoney, P., Buonanno, M., Welch, D., Adamson, C. S., Brenner, D. J., Noakes, C. and Wood, K. (2022) Far-UVC (222 nm) efficiently inactivates an airborne pathogen in a room-sized chamber. Sci. Reports 2022 121 12, 1–9. https://doi.org/10.1038/s41598-022-08462-z.

14. Finlayson, L., Barnard, I. R. M., McMillan, L., Ibbotson, S. H., Brown, C. T. A., Eadie, E. and Wood, K. (2022) Depth Penetration of Light into Skin as a Function of Wavelength from 200 to 1000 nm. Photochem. Photobiol. 98, 974–981. https://doi.org/10.1111/php.13550.

15. Eadie, E., Barnard, I. M. R., Ibbotson, S. H. and Wood, K. (2021) Extreme Exposure to Filtered Far-UVC: A Case Study. Photochem. Photobiol. 97, 527–531. https://doi.org/10.1111/php.13385.

16. Fukui, T., Niikura, T., Oda, T., Kumabe, Y., Ohashi, H., Sasaki, M., Igarashi, T., Kunisada, M., Yamano, N., Oe, K., Matsumoto, T., Matsushita, T., Hayashi, S., Nishigori, C. and Kuroda, R. (2020) Exploratory clinical trial on the safety and bactericidal effect of 222-nm ultraviolet C irradiation in healthy humans. PLoS One 15, e0235948. https://doi.org/10.1371/journal.pone.0235948.

17. Welch, D., Kleiman, N. J., Arden, P. C., Kuryla, C. L., Buonanno, M., Ponnaiya, B., Wu, X. and Brenner, D. J. (2022) No Evidence of Induced Skin Cancer or Other Skin Abnormalities after Long-Term (66 week) Chronic Exposure to 222-nm Far-UVC Radiation. Photochem. Photobiol. https://doi.org/10.1111/php.13656.

18. Yamano, N., Kunisada, M., Kaidzu, S., Sugihara, K., Nishiaki-Sawada, A., Ohashi, H., Yoshioka, A., Igarashi, T., Ohira, A., Tanito, M. and Nishigori, C. (2020) Long-term effects of 222 nm ultraviolet radiation C sterilizing lamps on mice susceptible to ultraviolet radiation. Photochem. Photobiol. 94, 853–862. https://doi.org/10.1111/php.13269.

19. Kaidzu, S., Sugihara, K., Sasaki, M., Nishiaki, A., Igarashi, T. and Tanito, M. (2019) Evaluation of acute corneal damage induced by 222-nm and 254-nm ultraviolet light in Sprague–Dawley rats. Free Radic. Res. 53. https://doi.org/10.1080/10715762.2019.1603378.

20. Kaidzu, S., Sugihara, K., Sasaki, M., Nishiaki, A., Ohashi, H., Igarashi, T. and Tanito, M. (2021) Re-Evaluation of Rat Corneal Damage by Short-Wavelength UV Revealed Extremely Less Hazardous Property of Far-UV-C†. Photochem. Photobiol. 97, 505–516. https://doi.org/10.1111/PHP.13419.

21. Kaidzu, S., Sugihara, K., Sasaki, M., Nishiaki, A., Ohashi, H., Igarashi, T. and Tanito, M. (2022) Safety evaluation of Far-UVC irradiation to epithelial basal cells in corneal limbus. In 41st American Society for Photobiology Biennial Meeting. Albuquerque.

22. Pitts, D. G. (1973) The ocular ultraviolet action spectrum and protection criteria. Health Phys. 25, 559–566. https://doi.org/10.1097/00004032-197312000-00002.

23. Chaney, E. K. and Sliney, D. H. (2005) Re-evaluation of the ultraviolet hazard action spectrum - The impact of spectral bandwidth. Health Phys. 89, 322–332. https://doi.org/10.1097/01.HP.0000164650.96261.9d.

24. Sugihara, K., Kaidzu, S., Sasaki, M., Ichioka, S., Takayanagi, Y., Shimizu, H., Sano, I., Hara, K. and Tanito, M. (2022) One-year Ocular Safety Observation of Workers and Estimations of Microorganism Inactivation Efficacy in the Room Irradiated with 222-nm Far Ultraviolet-C Lamps. Photochem. Photobiol. https://doi.org/10.1111/php.13710.

25. Wood, K., Wood, A., Peñaloza, C. and Eadie, E. (2022) Turn Up the Lights, Leave them On and Shine them All Around—Numerical Simulations Point the Way to more Efficient Use of Far-UVC Lights for the Inactivation of Airborne Coronavirus. Photochem. Photobiol. 98, 471–483. https://doi.org/10.1111/php.13523.

26. Blackie, C., Albou-Ganem, C. and Korb, D. (2012) Questionnaire assists in dry eye disease diagnosis. Four-question survey helps evaluate patients’ dry eye symptoms to improve screening. Oculat Surg. News Eur. Ed.

27. Schiffman, R. M., Christianson, M. D., Jacobsen, G., Hirsch, J. D. and Reis, B. L. (2000) Reliability and Validity of the Ocular Surface Disease Index. Arch. Ophthalmol. 118, 615–621. https://doi.org/10.1001/ARCHOPHT.118.5.615.

28. Duncan, M. A., Welch, D., Shuryak, I. and Brenner, D. J. (2022) Ocular and Facial Far-UVC Doses from Ceiling-Mounted 222 nm Far-UVC Fixtures. Photochem. Photobiol. https://doi.org/10.1111/php.13671.

29. First, M. W., Weker, R. A., Yasui, S. and Nardell, E. A. (2005) Monitoring human exposures to upper-room germicidal ultraviolet irradiation. J. Occup. Environ. Hyg. 2, 285–292. https://doi.org/10.1080/15459620590952224.

