## Supplementary figures and images for "222 nm Far-UVC from filtered Krypton-Chloride excimer lamps does not cause eye irritation when deployed in a simulated office environment"

### Supplementary Figure 1

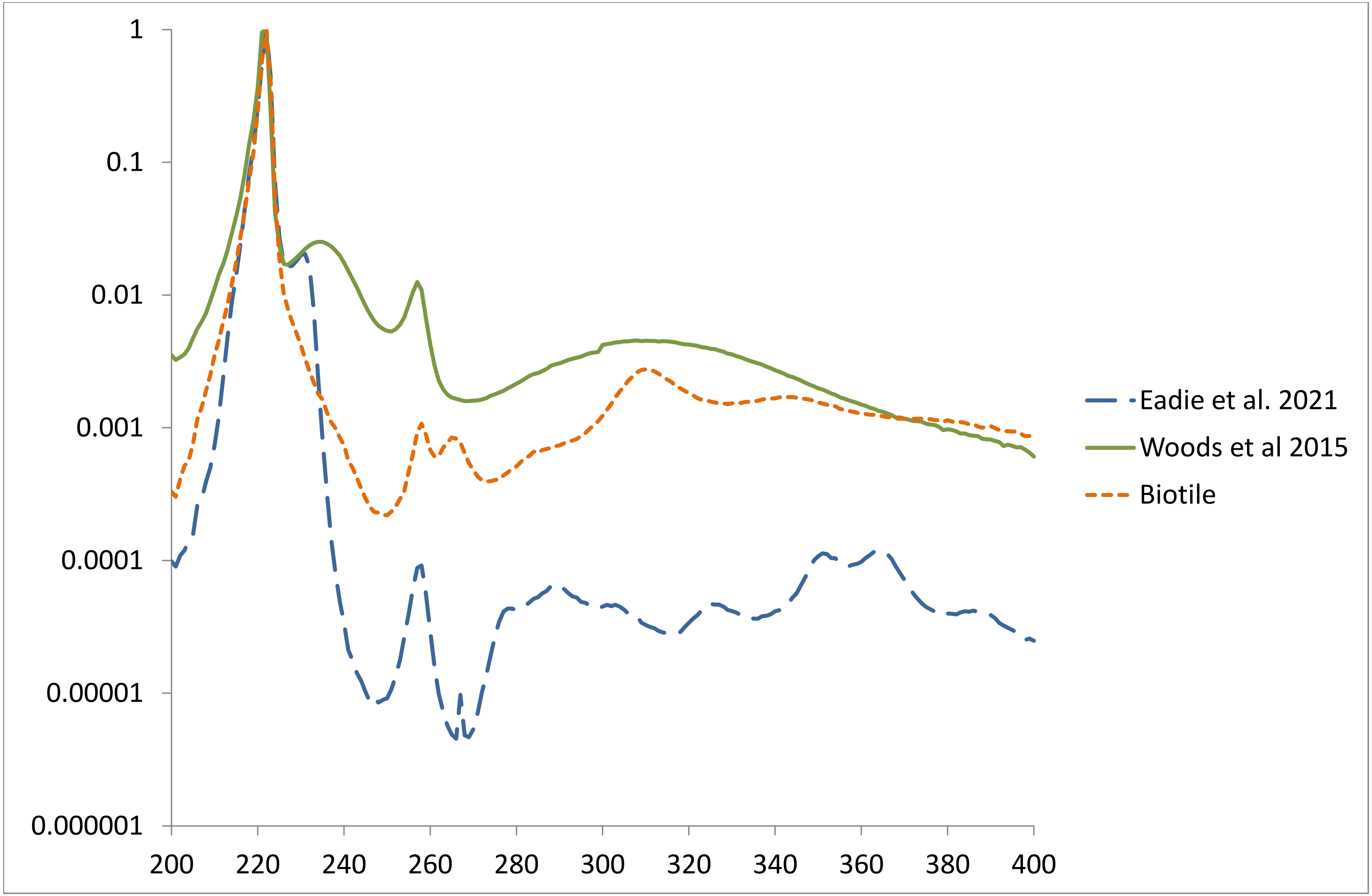
